# Perceived Gender Equitable Norms and Previous Tuberculosis Testing in Malawi: A Secondary Analysis of a Cluster-based Prevalence Survey

**DOI:** 10.1101/2025.04.23.25326292

**Authors:** Elizabeth Di Giacomo, Emily S. Nightingale, Peter MacPherson, Helena R.A. Feasey, Rebecca N. Soko, Vincent K. Phiri, Elizabeth L. Corbett, Katherine Horton

## Abstract

Substantial evidence demonstrates that men have a higher prevalence of tuberculosis (TB) and decreased use of TB services compared to women. Gender roles and norms contribute to these disparities by influencing social and structural determinants, as well as individual behaviours. In this analysis, we investigated attitudes towards gender equitable norms and TB testing behaviours amongst Malawian men and women participating in a prevalence survey conducted before a community-based TB active case finding trial in Blantyre. Perceptions of gender norms were captured through a modified version of the Gender Equitable Men Scale (GEMS). Gender inequitable views were prevalent among both men (56.1%) and women (55.8%). The association between a composite GEMS score and TB testing history was modelled using logistic regression, accounting for various sociodemographic covariates (age, sex, wealth quantile, education, and HIV status) (OR = 1.12, 95% CI: 0.88-1.42, p=0.373). Bivariate analysis demonstrated no notable confounding by any covariates and no strong effect modification. While GEMS score had no association with TB testing history among women, men with higher GEMS scores (less gender-equitable views) were more likely to have been tested for TB across age groups. These findings provide a basis for future investigation into the patterns and motives TB behaviours, particularly in older men. Tailored public health strategies may then be implemented to address this important population.

## Introduction

Tuberculosis (TB) remains a persistent global health and development challenge, with progress towards achieving the United Nations Sustainable Development Goal to end the global TB epidemic by 2030 far off track (1). An estimated 10.8 million people fell ill with TB in 2023, with 1.25 million deaths, making it the deadliest infectious disease among adults. From 2005-2019 there was a steady decrease in global TB incidence, however this trend reversed in 2020 and 2021 resulting in large reductions in notifications and associated increases in TB mortality (2). However, this fluctuation has slowly begun to stabilize (2).

Men are disproportionately affected by TB, accounting for 55% of all people who developed TB in 2023 and 52% of deaths (3). Prevalence surveys in high burden countries have shown that the underlying burden of undiagnosed TB is over twice as high among men as women, and comparisons of these findings to case notifications indicate that men are less likely than women to access timely diagnosis and treatment (4). The excess burden of disease and limited access to care among men may be attributed to social and structural determinants that increase men’s risk of disease, whilst limiting their ability to access diagnosis and treatment (5). These range from individual behaviours such as tobacco use and alcohol consumption, to social contact patterns and support networks, to social protections and institutional structures. Underlying many of these determinants are masculine social norms and expectations (4–9). Consequently, numerous calls to action are being made to address gender discrepancies in TB care, though subsequent action has been limited (10). Studies are beginning to uncover the specific challenges men face when seeking TB treatment, with recent work exploring the perceptions of masculinity and stigma surrounding TB among men (8, 9).

Gender norms have been defined as *“a subset of social norms that relate specifically to gender differences. They are informal, deeply entrenched and widely held beliefs about gender roles, power relations, standards or expectations that govern human behaviours and practices in a particular social context and at a particular time”* (11). They are considered important social determinants of health (12,13). Research has highlighted how masculine expectations of physical strength, stoicism, and control lead men to avoid admissions of illness and neglect TB symptoms until they become incapacitating, and how men deprioritise their own healthcare needs to fulfil roles as leaders and providers (5,9,14,15). We hypothesized that masculine expectations that affect health behaviours may be linked to attitudes towards gender norms, and that more equitable gender perceptions may be associated with improved access to TB services.

In this secondary analysis of data from a TB prevalence survey conducted in Malawi as part of a community cluster-randomised trial, we investigated the association between perceptions of gender norms and self-reported TB testing history using a validated tool to measure gender attitudes. We considered relevant covariates, and potential modification of this relationship by age and sex. A better understanding of the relationship between perspectives of gender norms and previous TB testing could identify new approaches and methods to reach men, in particular, with TB diagnostic services. This could result in more efficient public health strategies to improve diagnosis and treatment of individuals with TB and ultimately reduced transmission in the fight against TB, particularly among men as an underserved population.

## Materials and Methods

### Study Setting and Design

We conducted a secondary analysis of data from a community TB prevalence pre-intervention survey implemented before the active case finding cluster-randomised trial “Sustainable Community-wide Active Case Finding for Lung hEalth (SCALE)” (16,17). For the prevalence survey, households were randomly sampled from community neighbourhood clusters, and all adults (≥18 years) were invited to participate in a questionnaire and TB screening. A 20% random sub-sample of participants were additionally asked to complete an extended questionnaire that included questions on TB testing history, perceptions of gender equitable norms, healthcare utilization and social contacts mixing (Supplementary Figure 1). Criteria for inclusion were age of 18 years or older on the day of the interview; able to provide written or witnessed informed consent; intention to remain resident in Blantyre for at least 8 weeks following recruitment; and be willing to have radiological/microbiological screening for TB.

#### Measurements and procedures

For this analysis, the primary outcome was the proportion of participants who self-reported ever testing for TB. The primary outcome measure was participants’ response to two pre-intervention survey items on history of TB testing. The first item asked *“have you ever had a chest x-ray (CXR)”*. The second item asked *“have you ever given sputum for a medical test”*. Possible responses to each question were either “yes” or “no”.

The primary exposure was individual perception of gender equitable norms, measured using the Gender Equitable Men Scale (GEMS) (18). While the full GEMS consists of 42-items with domains covering violence, sexual relationships, reproductive health and disease prevention, and domestic chores and daily life (19), the SCALE Trial employed a modified 13-item version of the scale, available in both English and Chichewa (see S1 Table) as part of the pre-intervention phase. The scale is adaptable, and different versions have been used in a variety of settings (19). This particular adaptation was informed by previous work in Malawi investigating men’s gender attitudes and HIV risk, and a peer-delivered behavioural intervention on contraceptive uptake (20). As Pierotti’s (20)work added new items to the original GEMS scale, we received written permission to use these selected items in our study.

#### Covariates

The primary questionnaire captured data on participant’s age and sex, in addition to wealth indicators, education, marital status, employment, and HIV status. The self-assessed wealth index asked participants to position themselves on a six-step scale from poorest to richest, open to their interpretation and frame of reference. This question was taken from the Malawi 2016/2017 National Integrated Household Survey (21). The self-assessed wealth index was used as an individual’s attitude surrounding their wealth is likely more influential than objective measurements of durable assets in this context.

All participants were asked to report their HIV and antiretroviral therapy (ART) status, and were offered HIV testing. Individuals who self-reported a positive HIV test result, or ART use, or who tested positive were classified as HIV-positive.

#### Analysis

Analysis was conducted in Stata V.17.0 (22) and R Statistical software (v4.3.3) (23). We identified latent constructs underlying the 13 GEMS items through exploratory and confirmatory factor analysis (EFA and CFA). We undertook EFA with the principal factor method due to the non-normal distribution of responses to GEMS items (24). Any items with a uniqueness value >0.70 were excluded, as they signaled a high unique variation (25–27). We applied Promax rotation, and variables with high loadings were assessed to identify themes and inform the meaning of underlying latent factors (28). In assessing the number of factors to retain, we evaluated eigenvalues, scree plots, and performed a parallel analysis. We did not restrict our assessment to eigenvalues alone, as this technique has been cited as prone to both under- and over-factoring (29).

Using Cattell’s scree plot, we plotted the magnitude of eigenvalues from a reduced correlation matrix against the number of factors in descending order. We examined the plot for a substantial drop in the magnitude of eigenvalues and considered retaining the factor at and above this (30). Finally, we conducted a parallel analysis to benchmark the plotted points against predicted means of eigenvalues by 10,000 repeated sets of random data (24). We considered any points clearly above this line to represent factors that account for more variance than would be expected by chance (31).

Our CFA approach used standardised structural equation modelling (SEM) to construct a model of a set of predicted covariances between our latent variables and then test whether it is plausible when compared to the observed GEMS items (32). A post-estimation goodness-of-fit of the model was assessed by conducting a likelihood ratio test (LRT). The LRT chi-square results, the root mean squared error of approximation (RMSEA) (<0.08 acceptable), the comparative fit index (CFI) (>0.90 acceptable), and the Tucker-Lewis Index (TLI) (>0.90 acceptable) were assessed (26). Subsequently, factor scores were predicted using regression methods from the standardized SEM model (31), and merged into one composite score with equal weighting to form the primary exposure variable for the remainder of the analysis. Higher composite scores can be interpreted as more inequitable perspectives of gender norms, and vice versa.

To investigate the adjusted associations between exposures and outcomes, we first calculated stratum-specific odds ratios of self-reported TB testing history for each covariate with a corresponding 95% confidence interval, and significance level. We stratified GEMS score by TB testing history, and each covariate. We then constructed a logistic regression model of the principal association between the factor composite GEMS score and self-reported TB testing history, including additional co-variates based on *a priori* hypotheses and associations with GEMS items or TB health outcomes in similar settings (4,5,8,33–35). We incorporated a three-way interaction between the age, sex, and GEMS score to explore potential modification of the GEMS score association by these characteristics. Based on the fitted coefficients from the second model, we predicted the probability and 95% confidence interval for previous TB testing for illustrative values of age, sex, HIV status, and GEMS score.

### Ethics

Ethical approval for this secondary analysis of prevalence survey data (reference: 28700) – and for the SCALE Study (reference: 16228) – was obtained through the London School of Hygiene and Tropical Medicine’s Research Ethics Committee. In Malawi, ethics approval was obtained from the Malawian College of Medicine and Research Ethics Committee (COMREC, protocol number: P.12/18/2556). All participants provided written (or witnessed thumbprint, if illiterate) informed consent to participate in the parent study.

## Results

Of the 15,897 individuals enrolled in the SCALE Trial, 2,738 participants across 72 clusters completed the extended questionnaire containing the GEMS survey (Figure S1). This survey captured 2,278 unique households, with 1,720 households containing only a single participant (16). For this analysis of the study population who responded to the extended survey, 61% of participants were women (1,664) and 39% (1,074) men (Table 1), with a median age of 28 years (range 18 to 87 years). 98.8% (N=2,706) of participants had complete data for ever testing for TB, GEMS items, and all covariates of interest.

**Table 1:**
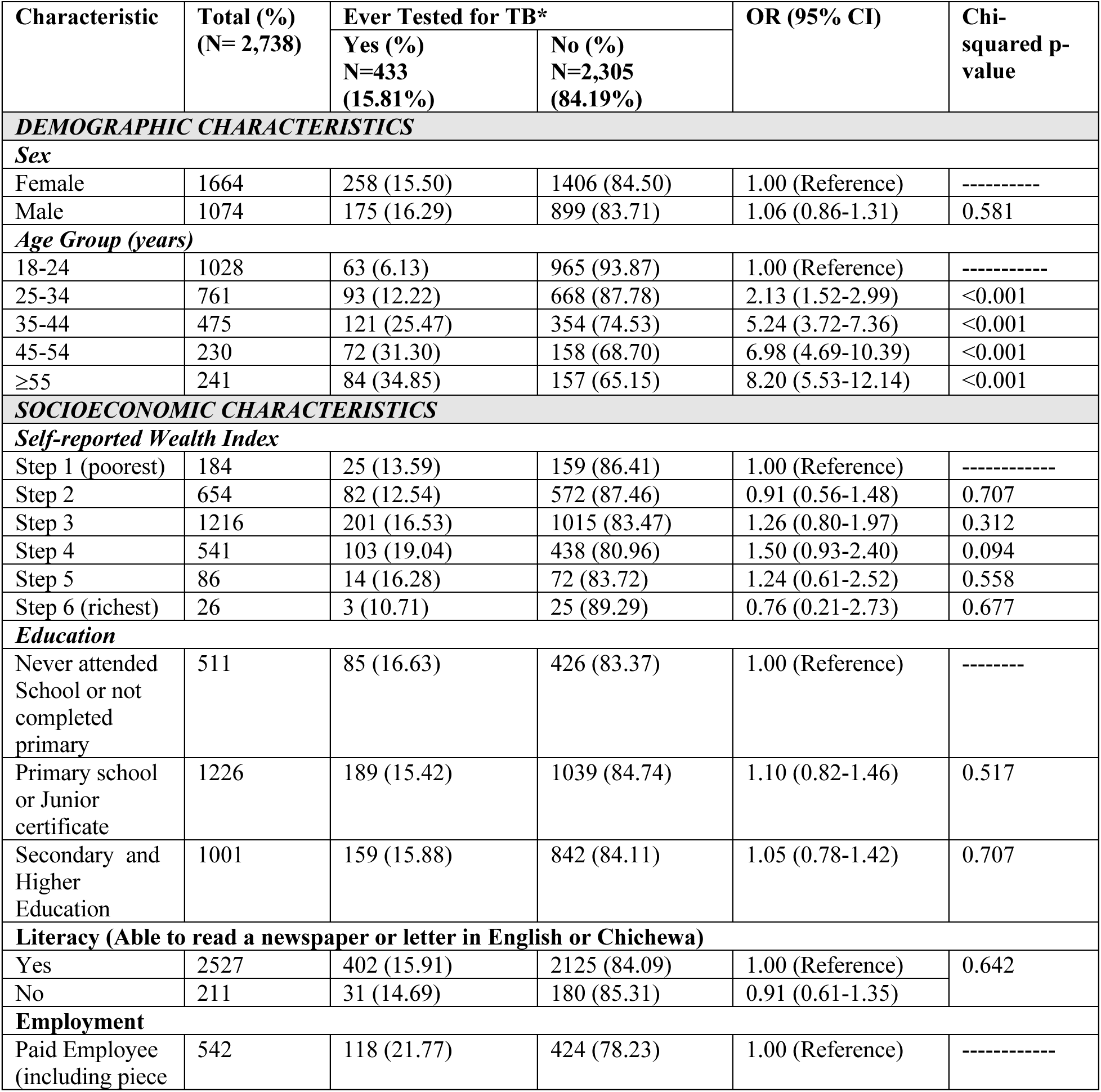
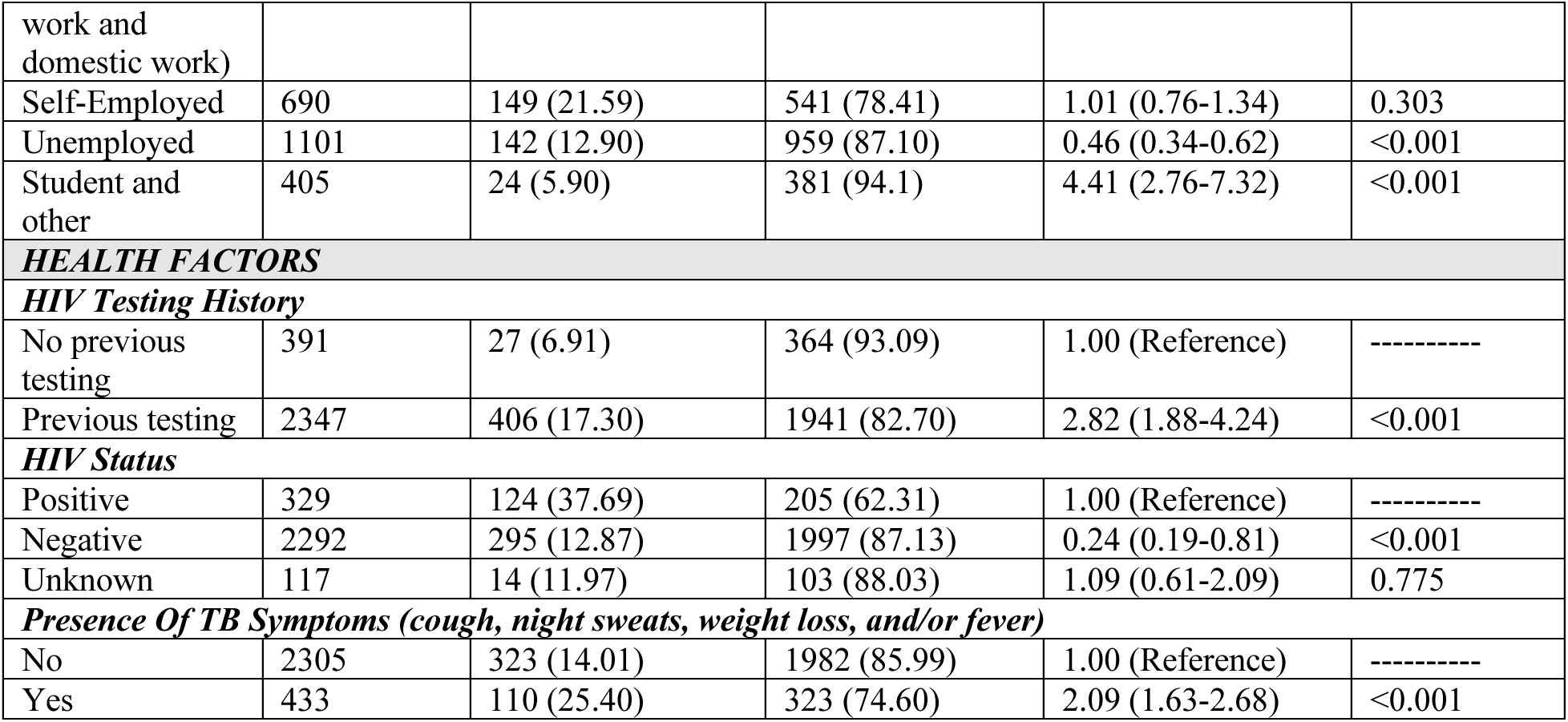
Characteristics of participants by tuberculosis testing history.

### Outcome Characteristics

15.8% (433/2,738) of the total population reported ever testing for TB (Table 1). 15.5% of women reported ever testing for TB, while 16.3% of men reported previously testing (Chi-square p=0.581). The proportion reporting ever testing was highest among individuals ages 35-44 years, with 25.5% having tested for TB in this age group. This age group had 5.24 times the odds of previous TB testing when compared to those in the lowest age group (95% CI: 3.72-7.36, p<0.001). Testing proportions were lowest among the youngest age group (17-24 years) with only 14.6% reporting having ever tested. Participant characteristics by HIV status are reported in Supplemental Table 1.

Amongst those who reported ever testing for TB, 93.8% also reported ever testing for HIV. This was slightly lower among those who did not report ever testing for TB, with 84.2% reporting also ever testing for HIV (Chi-square p<0.001). 37.7% of HIV-positive participants reported ever testing for TB, while only 12.9% of HIV-negative participants reported having done so (chi-square p<0.001). 25.4% of those who reported having any TB symptoms (cough, night sweats, unintentional weight loss, or fever) at the time of questionnaire administration also reported ever testing for TB. Interestingly, 14% of those who reported symptoms at the time of the questionnaire did not report previous testing for TB.

### Gender norms

Composite GEMS score factor and TB testing history are presented in Table 2. Higher GEMS composite scores can be interpreted as having more inequitable views of gender norms. Overall, there was no evidence of difference in GEMS scores distributions between participants who had, and had not, previously tested for TB (p=0.115). GEMS scores did differ slightly between HIV-positive and HIV-negative participants, with HIV-positive participants exhibiting more inequitable gender attitudes (p=0.044). The distribution of GEMS responses did not vary substantially between men and women (Figure 1). For all GEMS items except for Items 3, 9 and 10 (Supplementary Table 2), the majority of men and women reported strong agreement with the statement. Internal consistency does vary with adaption of the GEMS tool, however remained very high in the context of this study (Cronbach’s alpha = 0.88).

**Figure 1:**
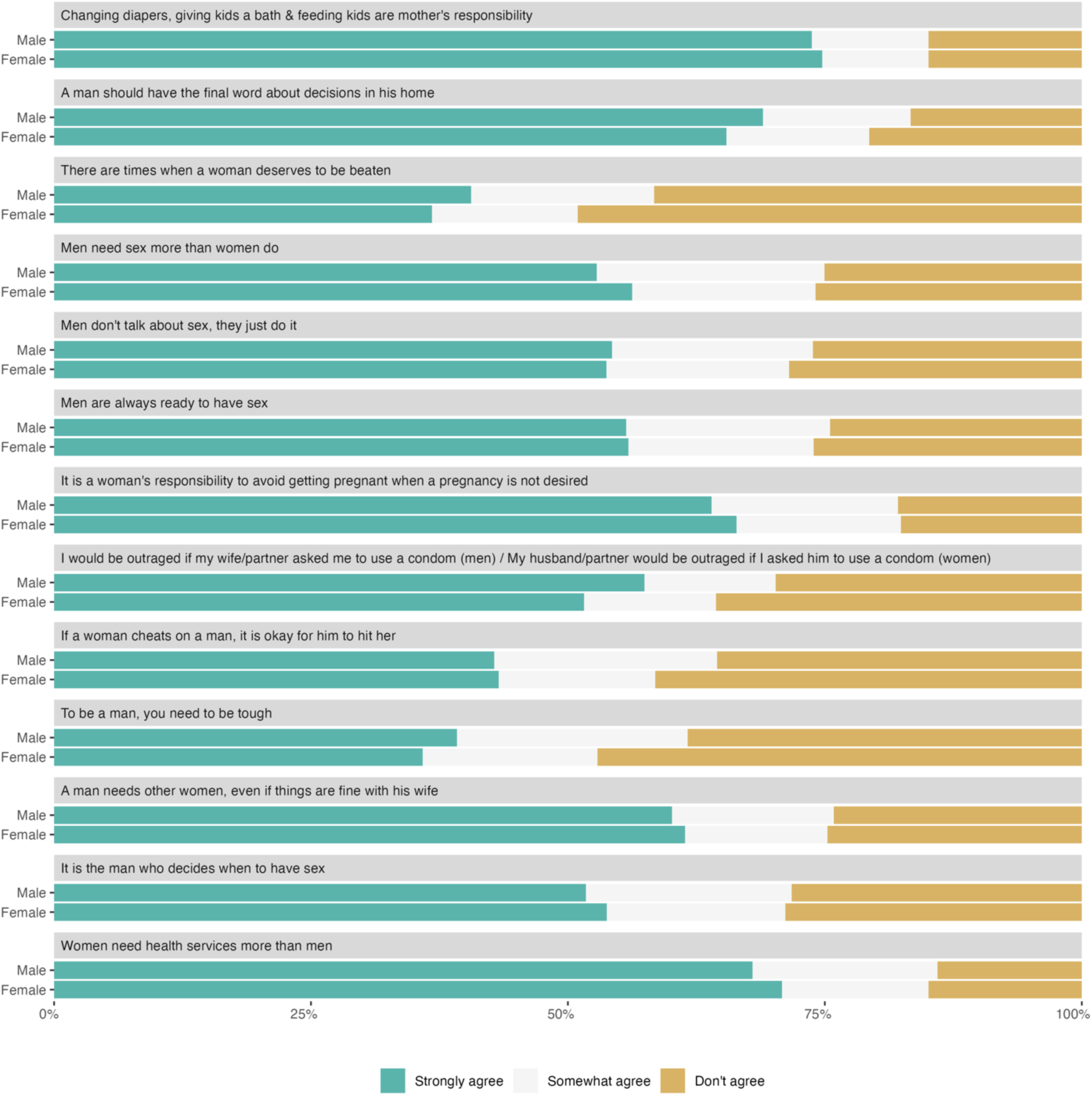
GEMS Responses, by Sex.

**Table 2:** Odds of prior TB testing, by age group, sex and HIV status.

| **HIV Status** | **Sex** | **N** | **Age Group** | **n (%)** | **OR for prior TB testing (95% CI)** | **p-value** |
| --- | --- | --- | --- | --- | --- | --- |
| *HIV Negative* | Female | 1364 | *17-24* | 514 (37.68) | **1.00 (Reference)** | ----- |
|  |  |  | *24-34* | 434 (31.82) | 1.46 (0.92-2.31) | 0.109 |
|  |  |  | *34-44* | 226 (16.57) | 3.21 (2.00-5.15) | <0.001 |
|  |  |  | *44-54* | 91 (6.67) | 4.23 (2.35-7.62) | <0.001 |
|  |  |  | *>55* | 99 (7.26) | 3.57 (1.98-6.44) | <0.001 |
|  | Male | 926 | *17-24* | 427 (46.11) | **1.00 (Reference)** | ----- |
|  |  |  | *24-34* | 238 (25.70) | 2.58 (1.43-4.65) | 0.002 |
|  |  |  | *34-44* | 118 (12.74) | 6.89 (3.78-12.56) | <0.001 |
|  |  |  | *44-54* | 58 (6.26) | 6.15 (2.92-12.95) | <0.001 |
|  |  |  | *>55* | 85 (9.18) | 13.53 (7.31-25.04) | <0.001 |
| *HIV Positive* | Female | 247 | *17-24* | 26 (10.53) | **1.00 (Reference)** | ----- |
|  |  |  | *24-34* | 64 (25.91) | 2.32 (0.70-7.65) | 0.166 |
|  |  |  | *34-44* | 92 (37.25) | 3.54 (1.13-11.10) | 0.031 |
|  |  |  | *44-54* | 46 (18.62) | 3.87 (1.15-13.06) | 0.029 |
|  |  |  | *>55* | 19 (7.69) | 6.11 (1.51-24.66) | 0.011 |
|  | Male | 79 | *17-24* | 2 (2.47) | **1.00 (Reference)** | ----- |
|  |  |  | *24-34* | 8 (9.88) | 0.32 (0.04-1.30) | 0.095 |
|  |  |  | 34-44 | 21 (25.93) | 0.40 (0.12-1.33) | 0.135 |
|  |  |  | 44-54 | 27 (33.33) | 0.51 (0.17-1.59) | 0.249 |
|  |  |  | >55 | 23 (28.40) | Omitted due to collinearity | ----- |

### Exploratory and Confirmatory Factor Analysis

Initial factoring of the 13 GEMS items revealed high uniqueness (>0.70) values for Item 1, Item 8 and Item 13 (diaper changing responsibility, condom usage, and women needing healthcare more, respectively). Cronbach’s alpha of the retained 10 items remained high at 0.87. Factor output from the polychoric correlation matrix using principal factor extraction with the remaining 10 variables demonstrated one factor with an eigenvalue >1. Examining the scree plot (S2 Figure), it was evident the point of inflection (“elbow”) of the curve is located at factor 2, where there is a steep rise in eigenvalue magnitude at factor 1. Factors one and two confidently exceed the threshold defined by the parallel analysis (S2 Figure). The resulting rotated factor loadings are given in S3 Table.

Questionnaire items loaded on to factor 1 appeared to relate to sexual autonomy and decisions. Conversely, factor 2 encompassed violence and physical toughness. They are termed “sexual factors” and “violence factors” in the analyses to follow. The internal consistency of both subscales was high, with alpha values of 0.84 and 0.72, respectively. The standardized SEM indicated good fit (RMSEA <0.05, CFI and TLI >0.90; Table S2).

#### Univariate Analysis

All variables were assessed for their potential association with GEMS composite scores and self-reported TB testing status. There was minimal difference in the odds of reporting having ever tested for TB between males and females (0.195 vs 0.184, respectively) with males having a 6% relative increase in odds compared to females, although these findings were not significant (OR = 1.06, 95% CI: 0.86-1.31, p=0.581). For age group, stratum-specific odds ratios for the outcome clearly demonstrated increasing odds of self-reporting ever testing for TB with increase in age group, with the oldest age group (55+ years of age) having 8.20 times the odds of reporting testing for TB compared to the 17–24-year-old age group (95% CI: 5.53-12.14, p<0.001). P-values provided strong evidence against the null hypothesis of no difference in odds (<0.001) for all age groups.

Those who were identified as positive for HIV had odds of reporting ever testing for TB nearly three-fold (OR: 2.82, 95% CI: 1.88-4.24, p<0.001) that of those who were defined as HIV negative. These results are supported by strong evidence against the null hypothesis of no difference in odds of reporting ever testing for TB between HIV groups.

#### Multivariate Analysis

In the logistic model that included GEMS score, TB testing history and all covariates, we did not identify a statistically significant association between participants’ GEMS score and their likelihood of having previously tested for TB (OR = 1.11, 95% CI: 0.87-1.43, p=0.396) (see S5 table). Increasing age group was strongly associated with prior TB testing (p<0.001), as was HIV status (p<0.001). Furthermore, a likelihood ratio test (LRT) for age provided evidence of a departure of linear trend (p=0.004), implying a potential non-linear relationship between age and previous TB testing history. A stratified analysis by HIV status demonstrated people living with HIV (PLHIV) had 50% higher odds of having previously tested for TB with each increasing unit of GEMS score, however this association was not statistically significant (p=0.088) (Table 2).

Inclusion of HIV status as a covariate in the model did not have a significant effect on the association between GEMS score and previous TB testing. The addition of HIV status decreases the odds ratio of previous TB testing from 1.17 (p=0.125) to only 1.12 (p=0.262).

The probability of previous TB testing among women increased with age (Figure 2). The probability of previous TB testing did not substantially vary by GEMS score in any age group for women.

**Figure 2:**
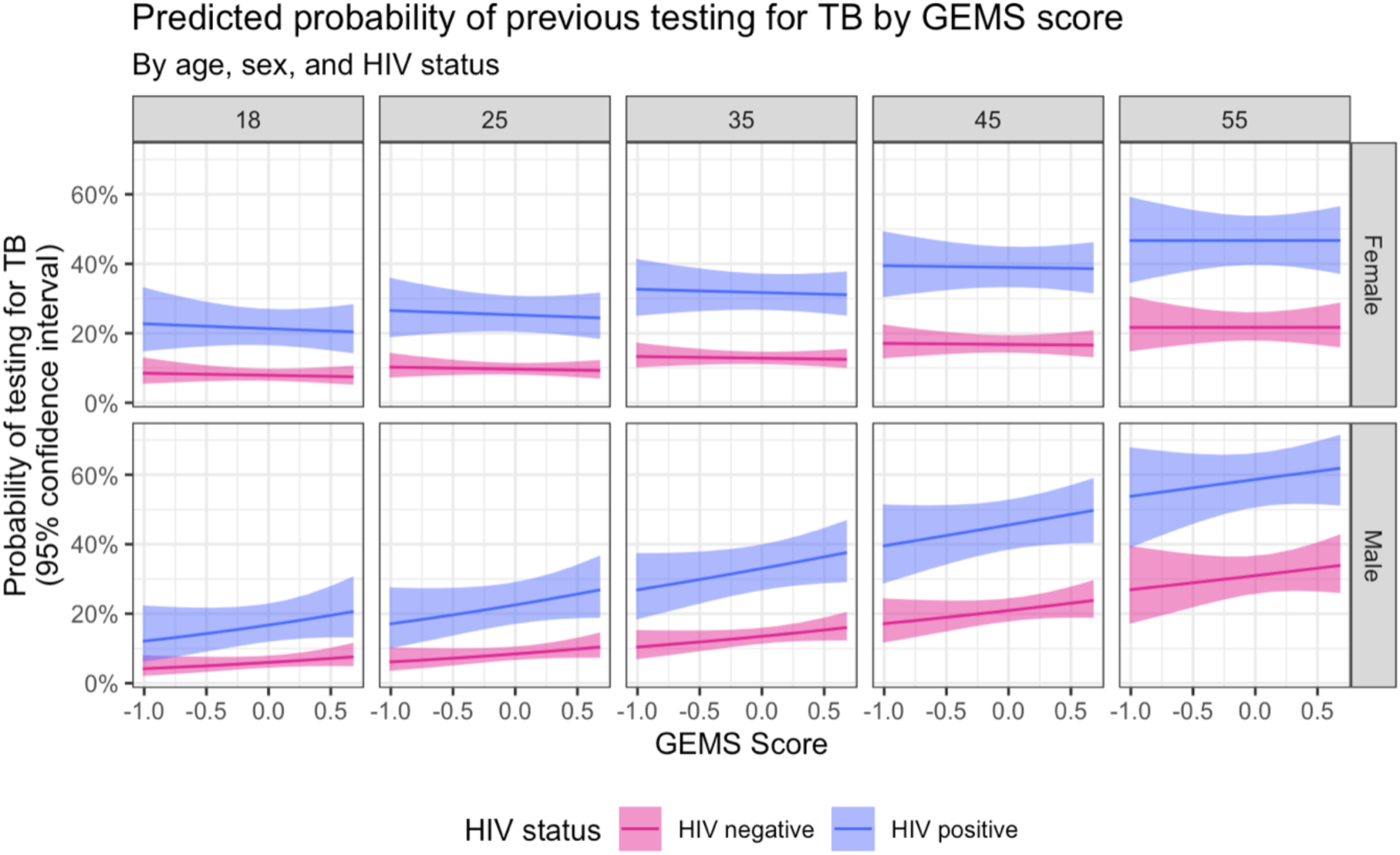
Predicted Probability of Ever Testing for TB, by GEMS Score, age, HIV status, and sex.

In men, the probability of reporting TB testing followed a similar distribution to women with respect to age. The probability of testing increased steadily across all age groups. However, there was more variability regarding GEMS score within age groups for men. In younger men (18 years of age), GEMS score had little effect on the probability of testing for TB. Among men 25 years old, the probability of testing for TB increased with GEMS score (less equitable gender perceptions). This pattern of increase was seen across all subsequent older age groups of men.

This trend is illustrated in Figure 2 through predicted probabilities at indicative ages for men and women, and a summary of the model fit is presented in Table S5. HIV-disaggregated trends are also shown in Figure 2.

## Discussion

Our analysis of perceptions of gender norms and self-reported TB testing behaviour found that negative attitudes towards gender equitable norms were prevalent in both men and women in Malawi. We did not detect a statistically significant association between perceptions of gender equity, as measured by GEMS, and the odds of self-reporting ever testing for TB. However, overall, we identified a potential effect of age on GEMS score and the probability of previous TB testing among men. For men, those with higher GEMS scores (less gender-equitable views) were more likely to have been tested for TB across age groups. It is notable that no associations between GEMS score and previous TB testing were noted among women.

To our knowledge, this is the first study to assess the impact of perceptions of gender equity and gender norms on a TB health outcome. The full GEMS tool has previously been used in Malawi to measure gender perspectives and assess the association with access to healthcare in central Malawi. This study found female gender, longer travel time to Lilongwe, and increased number of children in the household were independently associated with negative gender norm perspectives (33). Additional work exploring gender perspectives in Malawi has also been done using other measurement tools to assess gender perspectives on various outcomes (36–39). The GEMS score has been used in the Asia-Pacific region to investigate intimate partner violence, alcohol consumption and paternal parenting (40,41), as well as in India to explore women’s mistreatment during childbirth (42).

This study builds on previous investigations into TB prevalence data, which have highlighted a discrepancy between males and females in not only the frequency of TB testing but also engagement with available health services (primary care facilities, mobile digital x-ray vans, community-based symptom screening, and routine screening among people living with HIV and contacts of TB cases) (4,5,8,43). The latent factors extracted from this factor analysis agree with the themes and ideas expressed in existing literature exploring the drivers of sex differences in TB health-seeking behaviours. Sexual factors have been cited in previous qualitative work investigating drivers of the sex differences in health service access (6). Sexual factors were mentioned in the context of a man’s need for sex, and how a diagnosis of TB can impede opportunities for sexual activity in their relationship (6). Themes of physical violence were less explicitly stated in literature, however the GEMS items underpinning this latent factor are supported. The idea that toughness is inherent to masculinity is ubiquitous, and TB is understood by individuals to threaten or emasculate men (5,6).

The use of behavioural interventions to address gender attitudes have been previously investigated (44–46). The Intervention with Microfinance for AIDS and Gender Equity (IMAGE) study assessed the use of structural interventions, including training on gender roles, communication skills, and relationships. More progressive gender norms and a substantial reduction in intimate partner violence were reported (45). Community mobilization as an intervention has specifically been evaluated on GEMS responses, in the context of HIV in South Africa (46). The trial noted a statistically significant improvement in GEMS scores among men who received a theory-based, gender transformative intervention. This type of intervention aimed at shifting community gender norms may have far reaching effects for engagement with TB health services for men. A Cochrane review on active case finding highlighted that identification of cases is only part of the work to be done in promoting health service use for TB treatment.

Engendered stigma seen in HIV is compounded on to TB and contributes to discrimination, calling for community-based interventions to improve uptake (47). As a social construct, a platform for gender transformative education may be a valuable mechanism to promote community buy-in that is essential for shifts in gender attitudes.

Our results demonstrate an increase in history of previous TB testing with age and HIV status, which is consistent with accumulated testing opportunities over the life course and increased opportunities for TB testing for individuals in HIV care. This has been noted both in Malawi and other similar settings (43,48,49). Our study extended investigation into this relationship of age and TB testing history by considering both gender perspectives and sex and suggests that gender attitudes may affect testing behaviours among some men. The association between less gender-equitable attitudes and TB testing history may be associated with certain risk behaviours that increase the probability of contracting TB. In turn, this may increase their likelihood of being tested. However, it is notable that associations between GEMS score and TB testing history were consistent for HIV-positive and HIV-negative participants across age groups. Among women, gender attitudes did not appear to influence testing behaviours. We expect that attitudes towards gender equitable norms would have less impact on TB testing among women, given healthcare seeking behaviours are generally more acceptable for women. This may relate to women having more contact with health services, potentially for HIV screening or perinatal care. Gender attitudes would thus have an attenuated impact on care behaviours because it is more acceptable for women to receive this care.

This study was limited by binary structure of self-reported TB testing history, which may distill associations that exist with more specific elements of health service access and may be impacted by recall and social desirability bias. It may be worthwhile to evaluate this association in a rural setting, given that this study included only residents within the urban city of Blantyre. Important cultural differences may exist between rural and urban settings that change the nature of perceptions of gender norms as well as health access behaviours. Measuring GEMS scores at a single point in time and TB testing retrospectively services as a limitation, as attitudes towards both may have evolved over time. This calls for more data on how gender norms held in these settings may change over time, potentially through repeated surveys or longitudinal study designs relating gender norms to health outcomes. Finally, possibilities of accurate exposure ascertainment should be critically evaluated. As a complex and subjective social construct, questions surrounding how to best measure gender in this population and setting should be topics of ongoing discussion.

Although we found no statistically significant association between perceived gender equitable norms and self-reported TB testing history in this sample of participants living in Blantyre, Malawi, age appeared to influence GEMS score and the probability of previous TB testing, notably only among men. This study underscores the multifaceted nature of TB testing behaviours and wider healthcare access in Blantyre, Malawi. More nuanced research into this gender norm perspectives in the context of TB could inform gender transformative approaches to case identification and reach under-served populations, moving us closer to reaching EndTB targets in both Malawi and globally.

## Data Availability

The data that support the findings of this study are available on Open Science Framework (DOI 10.17605/OSF.IO/GY9JT). Due to ethical restrictions, some data has not been made publicly available.

## Supplementary Tables and Figures

**Supplementary Figure 1:**
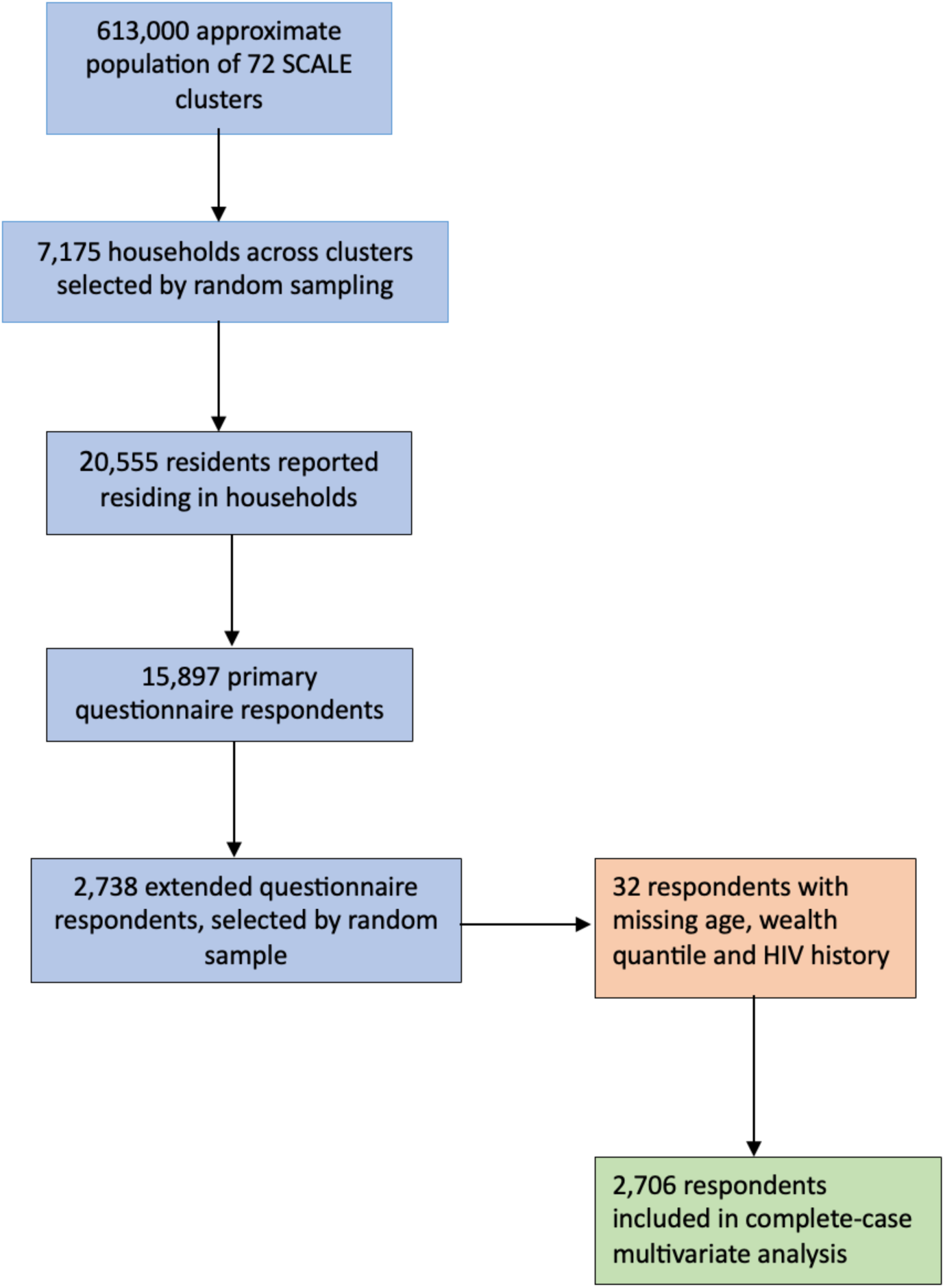
Flow Diagram of Study Population.

**Supplementary Figure 2:**
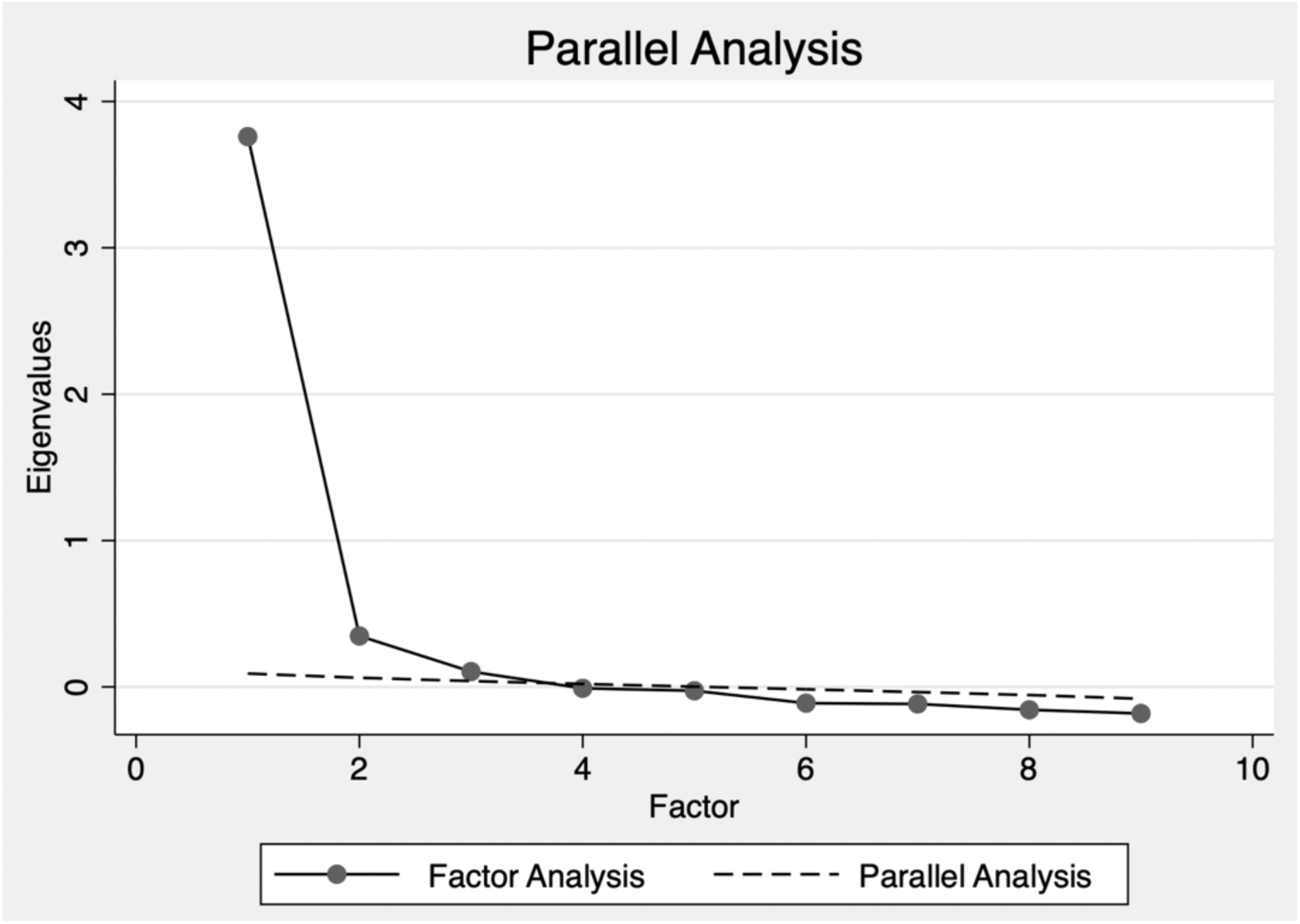
Scree Plot of Eigenvalues with Parallel Analysis*.

**Supplemental Table 1:** Characteristics of participants by HIV status.

| Characteristic | Total (%)<br>(N= 2,621) | HIV status |  | OR (95% CI) | Chi-squared p-value |
| --- | --- | --- | --- | --- | --- |
|  |  | Positive (%)<br>N=329<br>(12.55%) | Negative (%)<br>N=2,292<br>(87.45%) |  |  |
| DEMOGRAPHIC CHARACTERISTICS |  |  |  |  |  |
| Sex |  |  |  |  |  |
| Female | 1614 | 248 (15.37) | 1366 (84.63) | 1.00 (Reference) | <0.001 |
| Male | 1007 | 81 (8.04) | 926 (91.96) | 0.48 (0.37-0.63) |  |
| Age Group (years) |  |  |  |  |  |
| 18-24 | 969 | 28 (2.89) | 941 (97.11) | 1.00 (Reference) |  |
| 25-34 | 744 | 72 (9.68) | 672 (90.32) | 3.60 (2.30-5.63) | <0.001 |
| 35-44 | 457 | 113 (24.73) | 344 (75.27) | 11.04 (7.17-17.00) | <0.001 |
| 45-54 | 222 | 73 (32.88) | 149 (67.12) | 16.47 (10.30-26.31) | <0.001 |
| ≥55 | 226 | 42 (18.58) | 184 (81.42) | 7.67 (4.64-12.69) | <0.001 |
| SOCIOECONOMIC CHARACTERISTICS |  |  |  |  |  |
| Self-reported Wealth Index |  |  |  |  |  |
| Step 1 (poorest) | 179 | 36 (20.11) | 143 (79.89) | 1.00 (Reference) | ----- |
| Step 2 | 622 | 79 (12.7) | 543 (87.3) | 0.58 (0.37-0.89) | 0.013 |
| Step 3 | 1164 | 147 (12.63) | 1017 (87.37) | 0.57 (0.38-0.86) | 0.007 |
| Step 4 | 517 | 53 (10.25) | 464 (89.75) | 0.45 (0.29-0.72) | 0.001 |
| Step 5 | 85 | 9 (10.59) | 76 (89.41) | 0.47 (0.22-1.03) | 0.059 |
| Step 6 (richest) | 26 | 3 (11.54) | 23 (88.46) | 0.52 (0.14-1.82) | 0.305 |
| Education |  |  |  |  |  |
| Never attended School or not completed primary | 487 | 88 (18.07) | 399 (81.93) | 1.00 (Reference) | ----- |
| Primary school or Junior certificate | 1181 | 169 (14.31) | 1012 (85.69) | 0.76 (0.57-1.00) | 0.054 |
| Secondary and Higher Education | 953 | 72 (7.56) | 881 (92.44) | 0.37 (0.18-0.28) | <0.001 |
| Literacy (Able to read a newspaper or letter in English or Chichewa) |  |  |  |  |  |
| Yes | 2416 | 297 (12.29) | 2119 (87.71) | 1.00 (Reference) | 0.170 |
| No | 205 | 31 (15.12) | 173 (84.39) | 1.31 (0.89-1.96) |  |
| Employment |  |  |  |  |  |
| Paid Employee (including piece work and domestic work) | 516 | 74 (14.34) | 442 (85.66) | 1.00 (Reference) | ----- |
| Self-Employed | 663 | 111 (16.74) | 552 (83.26) | 1.20 (0.87-1.65) | 0.261 |
| Unemployed | 1066 | 138 (12.95) | 928 (87.05) | 0.89 (0.66-1.20) | 0.445 |
| Student and other | 376 | 6 (1.6) | 370 (98.4) | 0.10 (0.04-0.23) | <0.001 |

**Supplementary Table 2:** GEMS Scale Questions and Responses, by Sex.

| Item Number | Question* | Sex | Strongly Agree n (%) | Somewhat Agree n (%) | Do Not Agree n (%) |
| --- | --- | --- | --- | --- | --- |
| 1 | “Changing **diapers**, giving kids a bath & feeding kids are mother's responsibility.” | **Female** | 1244 (74.76) | 172 (10.34) | 248 (14.9) |
|  |  | **Male** | 792 (73.74) | 122 (11.36) | 160 (14.9) |
| 2 | “A man should have the final word about **decisions** in his home.” | **Female** | 1,089 (65.44) | 231 (13.88) | 344 (20.67) |
|  |  | **Male** | 741 (68.99) | 154 (14.34) | 179 (16.67) |
| 3 | “There are times when a woman deserves to be **beaten**.” | **Female** | 612 (36.78) | 236 (14.18) | 816 (49.04) |
|  |  | **Male** | 436 (40.60) | 191 (17.78) | 447 (41.62) |
| 4 | “Men need **sex more** than women do.” | **Female** | 936 (56.25) | 297 (17.85) | 431 (25.90) |
|  |  | **Male** | 567 (52.79) | 238 (22.16) | 269 (25.05) |
| 5 | “Men **don't talk** about sex, they just do it.” | **Female** | 894 (53.75) | 296 (17.79) | 474 (28.49) |
|  |  | **Male** | 583 (54.28) | 210 (19.55) | 281 (26.16) |
| 6 | “Men are **always ready** to have sex.” | **Female** | 930 (55.89) | 300 (18.03) | 434 (26.08) |
|  |  | **Male** | 598 (55.68) | 213 (19.83) | 263 (24.49) |
| 7 | “It is a woman's responsibility to avoid getting **pregnant** when a pregnancy is not desired.” | **Female** | 1105 (66.41) | 266 (15.99) | 293 (17.61) |
|  |  | **Male** | 687 (63.97) | 195 (18.16) | 192 (17.88) |
| 8 | “I would be outraged if my wife/partner asked me to use a **condom** (men) / My husband/partner would be outraged if I asked him to use a **condom** (women).” | **Female** | 858 (51.56) | 214 (12.86) | 592 (35.58) |
|  |  | **Male** | 617 (57.45) | 137 (12.76) | 320 (29.80) |
| 9 | “If a woman **cheats** on a man, it is okay for him to hit her.” | **Female** | 720 (43.27) | 254 (15.24) | 690 (41.47) |
|  |  | **Male** | 460 (42.83) | 233 (21.69) | 381 (35.47) |
| 10 | “To be a man, you need to be **tough**.” | **Female** | 597 (35.88) | 283 (17.01) | 784 (47.12) |
|  |  | **Male** | 421 (39.20) | 241 (22.44) | 412 (38.36) |
| 11 | “A man **needs other women**, even if things are fine with his wife.” | **Female** | 1022 (61.42) | 230 (13.82) | 412 (24.76) |
|  |  | **Male** | 646 (60.15) | 169 (15.74) | 259 (24.12) |
| 12 | “It is the man who **decides when** to have sex.” | **Female** | 895 (53.79) | 289 (17.37) | 480 (28.85) |
|  |  | **Male** | 556 (51.77) | 215 (20.02) | 303 (28.21) |
| 13 | “Women **need health services** more than men.” | **Female** | 1179 (70.85) | 237 (14.24) | 248 (14.90) |
|  |  | **Male** | 730 (67.97) | 193 (17.97) | 151 (14.06) |

**Supplementary Table 3:** Rotated Factor Loadings and CFA Goodness of Fit Statistics*.

| a. GEMS Items | Model 1: Two factor model (10 items) |  | Uniqueness |
| --- | --- | --- | --- |
|  | Sexual Autonomy and Decision-Making | Violence and Physical Toughness |  |
| It is the man who decides when to have sex | 0.423 |  | 0.688 |
| Men need sex more than women do | 0.739 |  | 0.462 |
| Men don't talk about sex, they just do it | 0.745 |  | 0.447 |
| Men are always ready to have sex | 0.738 |  | 0.435 |
| It is a woman's responsibility to avoid getting pregnant when a pregnancy is not desired | 0.462 |  | 0.707 |
| A man needs other women, even if things are fine with his wife | 0.330 |  | 0.650 |
| If a woman cheats on a man, it is okay for him to hit her |  | 0.680 | 0.703 |
| To be a man, you need to be tough |  | 0.754 | 0.730 |
| There are times when a woman deserves to be beaten |  | 0.363 | 0.683 |
| b. Fit indices (Robust) |  |  |  |
| Chi 2 (degrees of freedom) | <0.001 (26) |  |  |
| Comparative fit index (CFI) | 0.936 |  |  |
| Tucker-Lewis Index (TLI) | 0.911 |  |  |
| Root Mean Square Error of Approximation (RMSEA) | 0.050 |  |  |
| c. Factor correlations |  |  |  |
| Sexual Factors | 1.00 |  |  |
| Violence Factors | 0.884 | 1.00 |  |

**Supplementary Table 4:** Exploratory Obliquely Rotated Factor Loadings and Unique Variances (items 1, 8 and 13 excluded)

| **GEMS Description** | **Factor 1** | **Factor 2** | **Uniqueness\*** |
| --- | --- | --- | --- |
| There are times when a woman deserves to be beaten | 0.234 | **0.365** | 0.688 |
| Men need sex more than women do | **0.719** | 0.010 | 0.472 |
| Men don't talk about sex, they just do it | **0.750** | -0.001 | 0.440 |
| Men are always ready to have sex | **0.738** | 0.020 | 0.433 |
| It is a woman's responsibility to avoid getting pregnant when a pregnancy is not desired | **0.442** | 0.113 | 0.718 |
| If a woman cheats on a man, it is okay for him to hit her | 0.009 | **0.673** | 0.538 |
| To be a man, you need to be tough | -0.022 | **0.744** | 0.470 |
| A man needs other women, even if things are fine with his wife | **0.339** | 0.305 | 0.642 |
| It is the man who decides when to have sex | **0.422** | 0.345 | 0.491 |

**Supplementary Table 5:** Multivariable Regression Analysis Final Output.

|  | Characteristic | Adjusted OR | Wald Test p-value |
| --- | --- | --- | --- |
| Principal Association | GEMS Composite Score | 1.11 (0.87-1.43) | 0.396 |
| **Covariates** |  |  |  |
| Sex | Female | 1.00 (reference) | 0.146 |
|  | Male | 1.27 (0.92-1.78) |  |
| HIV Status | Negative | 1.00 (reference) | <0.001 |
|  | Positive | 2.69 (1.99-3.64) |  |
| Age Group | 17-24 | 1.00 (reference) | ----- |
|  | 25-34 | 1.56 (0.92-2.67) | 0.101 |
|  | 35-44 | 3.16 (1.86-5.38) | <0.001 |
|  | 45-54 | 3.41 (1.90-6.12) | <0.001 |
|  | ≥55 | 5.58 (3.09-10.10) | <0.001 |
| HIV Testing History | No | 1.00 (reference) | ----- |
|  | Yes | 2.75 (1.54-5.07) | <0.001 |
| Wealth Quantile | Step 1 | 1.00 (reference) | ----- |
|  | Step 2 | 1.03 (0.60-1.77) | 0.921 |
|  | Step 3 | 1.42 (0.86-2.37) | 0.186 |
|  | Step 4 | 1.67 (0.95-2.93) | 0.073 |
|  | Step 5 | 1.40 (0.60-3.24) | 0.438 |
|  | Step 6 | 0.36 (0.04-2.94) | 0.340 |
| Education | Never attended School or not completed primary | 1.00 (Reference) | ----- |
|  | Primary school leaving certificate | 1.30 (0.91-1.86) | 0.151 |
|  | Junior certificate of education | 0.92 (0.60-1.40) | 0.687 |
|  | Secondary with MSCE | 1.51 (1.01-2.26) | 0.044 |
|  | Higher Education | 1.36 (0.76-2.41) | 0.301 |
| Employment | Paid Employee | 1.00 (Reference) | ----- |
|  | Piece work (Ganyu) | 0.88 (0.51-1.52) | 0.723 |
|  | Paid domestic worker | 1.71 (0.41-7.09) | 0.508 |
|  | Self-Employed | 0.91 (0.63-1.32) | 0.718 |
|  | Unemployed | 0.70 (0.46-1.05) | 0.154 |
|  | Student | 4.62 (0.67-30.59) | 0.085 |
|  | Other | 0.49 (0.16-1.50) | 0.215 |
| Lost spouse to death | No | 1.00 (Reference) | ----- |
|  | Yes | 1.63 (1.08-2.46) | 0.020 |
| **Reliability Check Output** |  |  |  |
| Rho (p-value) |  | <0.001 (p=0.494) |  |

**Supplementary Table 6:** Interaction Model Coefficients.

| **Model Coefficient** | **Estimate** | **Standard Error** | **Z-Score** | **P-Value** |
| --- | --- | --- | --- | --- |
| **Intercept** | -3.028 | 0.194 | -15.586 | <0.001 |
| **GEMS Score** | -0.122 | 0.360 | -0.338 | 0.735 |
| **Sex** | -0.679 | 0.310 | -2.186 | 0.029 |
| **Age** | 0.032 | 0.005 | 6.480 | <0.001 |
| **GEMS Score : Sex** | 0.587 | 0.592 | 0.991 | 0.322 |
| **GEMS Score : Age** | 0.002 | 0.009 | 0.250 | 0.802 |
| **Sex : Age** | 0.021 | 0.007 | 2.843 | 0.004 |
| **GEMS Score : Sex : Age** | -0.007 | 0.014 | -0.509 | 0.611 |

## References

1. Kamalam DrS. Transforming Our World: The 2030 Agenda for Sustainable Development. Pondicherry Journal of Nursing. 2017;11(2).

2. World Health Organization. 2024 Global tuberculosis report. Geneva; 2024.

3. World Health Organization. Global tuberculosis report 2023 [Internet]. Geneva; 2023 [cited 2024 Oct 8]. Available from: https://www.who.int/teams/global-tuberculosis-programme/tb-reports/global-tuberculosis-report-2023

4. Horton KC, MacPherson P, Houben RMGJ, White RG, Corbett EL. Sex Differences in Tuberculosis Burden and Notifications in Low- and Middle-Income Countries: A Systematic Review and Meta-analysis. Vol. 13, PLoS Medicine. Public Library of Science; 2016.

5. Chikovore J, Pai M, Horton KC, Daftary A, Kumwenda MK, Hart G, et al. Missing men with tuberculosis: The need to address structural influences and implement targeted and multidimensional interventions. Vol. 5, BMJ Global Health. BMJ Publishing Group; 2020.

6. Miller C, Huston J, Samu L, Mfinanga S, Hopewell P, Fair E. “It makes the patient’s spirit weaker”: Tuberculosis stigma and gender interaction in Dar es Salaam, Tanzania. International Journal of Tuberculosis and Lung Disease. 2017;21.

7. Feasey HRA, Burke RM, Nliwasa M, Chaisson LH, Golub JE, Naufal F, et al. Do community-based active case-finding interventions have indirect impacts on wider TB case detection and determinants of subsequent TB testing behaviour? A systematic review. PLOS Global Public Health. 2021 Dec 8;1(12):e0000088.

8. Chikovore J, Hart G, Kumwenda M, Chipungu G, Desmond N, Corbett EL. TB and HIV stigma compounded by threatened masculinity: Implications for TB health-care seeking in Malawi. International Journal of Tuberculosis and Lung Disease. 2017 Nov 1;21:S26–33.

9. Chikovore J, Hart G, Kumwenda M, Chipungu GA, Desmond N, Corbett L. Control, struggle, and emergent masculinities: A qualitative study of men’s care-seeking determinants for chronic cough and tuberculosis symptoms in Blantyre, Malawi. BMC Public Health. 2014;14(1).

10. Ringwald B, Mwiine AA, Chikovore J, Makanda G, Amoah-Larbi J, Millington KA, et al. Ending TB means responding to socially produced vulnerabilities of all genders. Vol. 8, BMJ Global Health. 2023.

11. UNICEF. Technical Note on Gender Norms. 2020.

12. Courtenay WH. Constructions of masculinity and their influence on men’s well-being: A theory of gender and health. Soc Sci Med. 2000;50(10).

13. European Institute for Gender Equality. Gender Norms [Internet]. 2016 [cited 2023 Sep 1]. Available from: https://eige.europa.eu/publications-resources/thesaurus/terms/1288?language_content_entity=en

14. Chikovore J, Hart G, Kumwenda M, Chipungu GA, Corbett L. “For a mere cough, men must just chew Conjex, gain strength, and continue working”: The provider construction and tuberculosis care-seeking implications in Blantyre, Malawi. Glob Health Action. 2015;8(1).

15. Daniels J, Medina-Marino A, Glockner K, Grew E, Ngcelwane N, Kipp A. Masculinity, resources, and retention in care: South African men’s behaviors and experiences while engaged in TB care and treatment. Soc Sci Med. 2021;270.

16. Nightingale ES, Feasey HRA, Khundi M. Community-level variation in TB testing history: analysis of a prevalence survey in Blantyre, Malawi. MedRxiv [Internet]. 2023 May 2 [cited 2023 Sep 20]; Available from: https://www.medrxiv.org/content/10.1101/2023.04.28.23289249v1

17. Wellcome Senior Research Fellowships. Sustainable Community Action for Lung hEalth (SCALE): a cluster randomised trial in Blantyre, Malawi [Internet]. 2016 [cited 2023 Sep 1]. Available from: https://wellcome.org/grant-funding/people-and-projects/grants-awarded/sustainable-community-action-for-lung-health

18. Pulerwitz J, Barker G. Measuring attitudes toward gender norms among young men in Brazil: Development and psychometric evaluation of the GEM scale. Men Masc. 2008;10(3).

19. Nanda G. Compendium of Gender Scales [Internet]. Washington, DC; 2011 Sep [cited 2023 Sep 27]. Available from: www.c-changeproject.org

20. Pierotti SR. Men’s Gender Attitudes and HIV Risk in Urban Malawi. [Ann Arbor]: University of Michigan; 2013.

21. The World Bank Group. Fourth Integrated Household Survey 2016-2017 (IHS4)/ Malawi Living Standards Measurement Survey - Integrated Surveys on Agriculture 16/17 / Malawi IHS3 - Year: 3 [Internet]. 2021 Apr [cited 2023 Sep 1]. Available from: https://microdata.worldbank.org/index.php/catalog/2936/study-description

22. StataCorp. Stata Statistical Software: Release 17. College Station, TX: StataCorp LLC; 2021.

23. R Core Team. R: A language and environment for statistical computing. Vienna, Austria: R Foundation for Statistical Computing; 2024.

24. Fabrigar LR, MacCallum RC, Wegener DT, Strahan EJ. Evaluating the use of exploratory factor analysis in psychological research. Vol. 4, Psychological Methods. 1999.

25. Johnson RA, Wichern DW. Applied Multivariate Statistical Analysis.: Pearson Prentice Hall. Pearson Prentice Hall. 2007.

26. Streiner DL. Building a better model: An introduction to structural equation modelling. Canadian Journal of Psychiatry. 2006;51(5).

27. Cattell RB, Vogelmann S. A comprehensive trial of the scree and kg criteria for determining the number of factors. Multivariate Behav Res. 1977;12(3).

28. Hendrickson AE, White PO. PROMAX: A QUICK METHOD FOR ROTATION TO OBLIQUE SIMPLE STRUCTURE. British Journal of Statistical Psychology. 1964;17(1).

29. Zwick WR, Velicer WF. Comparison of Five Rules for Determining the Number of Components to Retain. Psychol Bull. 1986;99(3).

30. Cattell RB. The scree test for the number of factors. Multivariate Behav Res. 1966;1(2).

31. Brown T. Confirmatory Factor Analysis for Applied Research, Second Edition. Guilford Publications. 2015.

32. Franke GR, Mueller RO. Basic Principles of Structural Equation Modeling: An Introduction to LISREL and EQS. Journal of Marketing Research. 1996;33(2).

33. Azad AD, Charles AG, Ding Q, Trickey AW, Wren SM. The gender gap and healthcare: associations between gender roles and factors affecting healthcare access in Central Malawi, June–August 2017. Archives of Public Health. 2020;78(1).

34. Berhan A, Almaw A, Solomon Y, Legese B, Damtie S, Erkihun M, et al. Tuberculosis Treatment Outcome and Associated Factors Among Tuberculosis Patients Linked to Tuberculosis Treatment Clinics in Ethiopia, 2023: A Multi-Center Retrospective Study. Infect Drug Resist. 2023;16.

35. Agazhu HW, Assefa ZM, Beshir MT, Tadesse H, Mengstie AS. Treatment outcomes and associated factors among tuberculosis patients attending Gurage Zone Public Hospital, Southern Nations, Nationalities, and People’s Region, Ethiopia: an institution-based cross-sectional study. Front Med (Lausanne). 2023;10.

36. Mphangwe W, Nolan A, Vallieres F, Finn M. How do gender norms contribute to stunting in Ntchisi District, Malawi? A qualitative study. medRxiv. 2023;

37. Mudege NN, Mdege N, Abidin PE, Bhatasara S. The role of gender norms in access to agricultural training in Chikwawa and Phalombe, Malawi. Gender, Place and Culture. 2017;24(12).

38. Mudege NN, Kapalasa E, Chevo T, Nyekanyeka T, Demo P. Gender norms and the marketing of seeds and ware potatoes in Malawi. Journal of Gender, Agriculture and Food Security. 2015;1(2).

39. Mudege NN, Chevo T, Nyekanyeka T, Kapalasa E, Demo P. Gender Norms and Access to Extension Services and Training among Potato Farmers in Dedza and Ntcheu in Malawi. Journal of Agricultural Education and Extension. 2016;22(3).

40. Laslett AM, Graham K, Wilson IM, Kuntsche S, Fulu E, Jewkes R, et al. Does drinking modify the relationship between men’s gender-inequitable attitudes and their perpetration of intimate partner violence? A meta-analysis of surveys of men from seven countries in the Asia Pacific region. Vol. 116, Addiction. 2021.

41. Laslett AM, Kuntsche S, Wilson IM, Taft A, Fulu E, Jewkes R, et al. The relationship between fathers’ heavy episodic drinking and fathering involvement in five Asia-Pacific countries: An individual participant data meta-analysis. Alcohol Clin Exp Res. 2022;46(12).

42. Diamond-Smith N, Treleaven E, Murthy N, Sudhinaraset M. Women’s empowerment and experiences of mistreatment during childbirth in facilities in Lucknow, India: Results from a cross-sectional study. BMC Pregnancy Childbirth. 2017;17.

43. Feasey HRA, Khundi McEwen, Nzawa Soko R, Nightingale Emily. Prevalence of Bacteriologically-Confirmed Tuberculosis in Urban Blantyre, Malawi 2019-20: Substantial Decline Compared to 2013-14 National Survey. MedRxiv [Internet]. 2023 Apr 23 [cited 2023 Sep 27]; Available from: 10.1101/2023.04.20.23288872

44. Pulerwitz J, Hughes L, Mehta M, Kidanu A, Verani F, Tewolde S. Changing gender norms and reducing intimate partner violence: Results from a quasi-experimental intervention study with young men in Ethiopia. Am J Public Health. 2015;105(1).

45. Pronyk PM, Hargreaves JR, Kim JC, Morison LA, Phetla G, Watts C, et al. Effect of a structural intervention for the prevention of intimate-partner violence and HIV in rural South Africa: a cluster randomised trial. Lancet. 2006;368(9551).

46. Pettifor A, Lippman SA, Gottert A, Suchindran CM, Selin A, Peacock D, et al. Community mobilization to modify harmful gender norms and reduce HIV risk: results from a community cluster randomized trial in South Africa. J Int AIDS Soc. 2018;21(7).

47. Medley N, Taylor M, van Wyk SS, Oliver S. Community views on active case finding for tuberculosis in low- and middle-income countries: a qualitative evidence synthesis. Cochrane Database of Systematic Reviews. 2021;2021(3).

48. Moyo S, Ismail F, Van der Walt M, Ismail N, Mkhondo N, Dlamini S, et al. Prevalence of bacteriologically confirmed pulmonary tuberculosis in South Africa, 2017–19: a multistage, cluster-based, cross-sectional survey. Lancet Infect Dis. 2022 Aug 1;22(8):1172–80.

49. Feasey HRA, Corbett EL, Nliwasa M, Mair L, Divala TH, Kamchedzera W, et al. Tuberculosis diagnosis cascade in Blantyre, Malawi: a prospective cohort study. BMC Infect Dis. 2021;21(1).

